# Effect of Shenfu Injection on Myocardial Injury After Primary PCI for STEMI: The RESTORE Trial

**DOI:** 10.1101/2024.07.01.24309807

**Authors:** Xiao Wang, Zeyuan Fan, Jianjun Jiang, Jincheng Guo, Zhifang Wang, Likun Ma, Ruili He, Guohai Su, Hongtao Liu, Delu Yin, Guoan Zhao, Yingying Guo, Meixia Shang, Xinliang Ma, Shaoping Nie

**Affiliations:** Cardiometabolic Medicine Center, Fuwai Hospital, National Center for Cardiovascular Diseases, Chinese Academy of Medical Sciences and Peking Union Medical College, Beijing, China; Center for Coronary Artery Disease, Division of Cardiology, Beijing Anzhen Hospital, Capital Medical University, Beijing, China; Department of Cardiovascular Diseases, Civil Aviation General Hospital, Civil Aviation Clinical Medical College of Peking University, Beijing, China; Department of Cardiology, Taizhou Hospital of Zhejiang Province affiliated to Wenzhou Medical University, Linhai, Zhejiang, China; Department of Cardiology, Beijing Luhe Hospital, Capital Medical University, Beijing, China; Department of Cardiology, Xinxiang Central Hospital, The Fourth Clinical College of Xinxiang Medical University, Xinxiang, Henan, China; Department of Cardiology, the First Affiliated Hospital of USTC, Division of Life Sciences and Medicine, University of Science and Technology of China, Hefei, Anhui, China; Department of Cardiology, Huaihe Hospital of Henan University, Kaifeng, China; Department of Cardiology, Central Hospital Affiliated to Shandong First Medical University, Jinan, Shandong, China; Department of Cardiology, Shenzhen Longhua District Central Hospital, The Affiliated Central Hospital of Shenzhen Longhua District, Guangdong Medical University, Shenzhen, China; Department of Cardiology, The First Hospital of Lianyungang, Xuzhou Medical University, Lianyungang, China; Department of Cardiology, The First Affiliated Hospital of Xinxiang Medical University, Weihui, China; Department of Biostatistics, Peking University First Hospital, Beijing, China; Department of Emergency Medicine, Thomas Jefferson University, Philadelphia, PA, USA

**Keywords:** cardiac magnetic resonance imaging, myocardial infarction, myocardial injury, primary percutaneous coronary intervention, Shenfu injection

## Abstract

**BACKGROUND:** Shenfu injection, as a traditional Chinese medicine, can alleviate reperfusion injury after ST-segment elevation myocardial infarction (STEMI) through multiple pharmacologic effects. This trial aimed to evaluate the effect of Shenfu injection on myocardial injury in STEMI patients undergoing primary percutaneous coronary intervention (PCI).

**METHODS:** This was a multicenter, randomized, double-blind, parallel-group, placebo-controlled trial. First-time anterior STEMI patients undergoing primary PCI within 12 hours of symptom onset due to a proximal or mid left anterior descending artery occlusion were randomized 1:1 to receive either intravenous Shenfu injection or placebo before reperfusion and followed by once a day until 5 days after primary PCI. The primary endpoint was infarct size by cardiac magnetic resonance (CMR) at 5 days after randomization.

**RESULTS:** A total of 295 patients were randomized with evaluable CMR in 273 patients. Infarct size (37.4±14.1% vs 37.5±14.5%; effect size −0.04%, 95% confidence interval: −3.45, 3.37; P=0.982) did not differ between the Shenfu injection and placebo groups. This was true for other CMR parameters. The area under curve for creatine kinase-myocardial band did not differ between groups. The incidences of thrombolysis in myocardial infarction (TIMI) flow grade 3 (94.8% vs 97.1%, P=0.324), TIMI myocardial perfusion grade 3 (91.7% vs 92.1%, P=0.914), and ST-segment resolution ≥70% (25.4% vs 26.1%, P=0.906), were also similar between groups. Adverse events were evenly distributed across groups.

**CONCLUSIONS:** For patients with anterior STEMI undergoing primary PCI, administration of Shenfu injection was safe but did not reduce infarct size by CMR.

**REGISTRATION:** URL: https://www.clinicaltrials.gov; Unique identifier: NCT04493840.

**Clinical Perspective:** *What Is New?:*

- Among 295 anterior ST-segment elevation myocardial infarction patients randomized to intravenous Shenfu injection or placebo, the primary analysis demonstrated that the infarct size revealed by cardiac magnetic resonance was 37.4 % in the Shenfu injection group and 37.5% in the placebo group, a difference that was not statistically significant.
- For patients with anterior ST-segment elevation myocardial infarction patients undergoing primary percutaneous coronary intervention, administration of Shenfu injection was safe and well tolerated.

*What Are the Clinical Implications?:*

- Cardiovascular clinicians need to realize that successful cardioprotective therapies may require synergistic multitarget approaches.
- Future clinical trials are required to investigate the use of additive cardioprotective strategies in patients with large infarcts or severe hemodynamic alterations (eg. concomitant heart failure or cardiogenic shock).

## INTRODUCTION

Despite advances in reperfusion strategies and concomitant antithrombotic therapy, the morbidity and mortality of acute ST-segment elevation myocardial infarction (STEMI) remain substantial^1^. Reperfusion therapy rescues the ischemic myocardium but paradoxically induces myocardial injury^2–4^. Multiple cardioprotective therapies have been proposed to reduce infarct size but showed inconsistent clinical efficacy^5–11^. Specifically, multifaceted pathophysiological factors (endothelial dysfunction, microvascular obstruction, calcium overload, oxidative stress, inflammation, and mitochondrial dysfunction) were involved in the process of reperfusion injury^2, 12–14^. As such, the most promising approach to cardioprotection may be to combine multitarget therapy with synergistic cardioprotective effect.

Shenfu injection, a component-based Chinese medicine with “Yang”-restoring and “Qi”-tonifying property, is exacted from ginseng (Panax; family: Araliaceae) and aconite (Radix aconiti lateralis preparata, Aconitum carmichaeli Debx; family: Ranunculaceae)^15, 16^ with ginsenosides and aconite alkaloids as the main active ingredients, containing 1 mg/ml Panax ginseng C.A. Mey and 2 mg/ml Aconitum Carmichaeli Debeaux^17, 18^. Shenfu injection exerts synergistic cardioprotective effect by forming a complex pharmacologic network directed to distinct signaling pathways within the cardiomyocyte, and it could improve the efficacy by the combination of multi-components and multitargets^17, 19^. In our pilot study, we demonstrated the feasibility and safety of Shenfu injection after primary percutaneous coronary intervention (PCI) for STEMI^20^. Consequently, we designed the RESTORE (Randomized Evaluation of Shenfu Injection to Reduce Myocardial Injury) trial powered to assess the effects of Shenfu injection on myocardial infarct size as determined by cardiac magnetic resonance (CMR) imaging in patients with STEMI undergoing primary PCI.

## METHODS

### Study design

The RESTORE trial was an investigator-initiated, multicenter, randomized, double-blind, parallel-group, placebo-controlled trial to evaluate the effect of Shenfu injection compared with placebo on myocardial injury in patients with a first-time anterior STEMI following successful primary PCI, conducted in accordance with the Helsinki Declaration. This trial was approved by the Institutional Review Board of Beijing Anzhen Hospital, Capital Medical University (2019013) and the institutional review committee at each clinical center. All participants provided written informed consent prior to randomization. This trial was registered at ClinicalTrials.gov (NCT04493840).

A more detailed protocol has been previously reported^21^. Briefly, patients aged 18-75 years, with first-time anterior STEMI undergoing primary PCI with the time from onset of ischemic symptom to the time of initial balloon inflation ≤ 12 hours were eligible for randomization. Further angiographic criteria included the presence of left anterior descending branch (LAD) occlusion in proximal or middle segment with pre-PCI thrombolysis in myocardial infarction (TIMI) flow 0 or 1. Key exclusion criteria were cardiogenic shock, serious heart failure, malignant ventricular arrhythmia, mechanical complications, prior myocardial infarction, PCI or bypass surgery, and known contraindication for CMR imaging. Full eligibility and exclusion criteria are shown in Table S1. The participants were enrolled between July 2020 and May 2023 in eleven regional cardiac centers in China.

### Randomization and blindness

Patients were randomly assigned 1:1 to either Shenfu injection or matching placebo by using an Interactive Web Response System (IWRS) before primary PCI and the randomization sequence was performed in blocks of 4. The IWRS was maintained by a third-party of the Institute of Clinical Research, Peking University. The randomization number assigned by IWRS automatically corresponded with a unique medication number of research drugs (Shenfu injection or placebo).

The Shenfu injection and placebo were provided by the collaborator, China Resources Sanjiu Medical & Pharmaceutical Co, Ltd, and packaged based on the randomization allocation list and the blinded principle. The study drug Shenfu injection was a colored transparent liquid, while the placebo 5% glucose injection was a colorless transparent liquid. For reducing the bias, a disposable lucifugal infusion bag and apparatus was used during the infusion to efficiently cover the study drug and matched placebo. To maintain the double-blind design, only the designated unblinded medical professional knows the group information. The injection was prepared by the medical professional according to the dosage of the study drug in a separate room. All the medical professionals followed the blind and mask standard operating procedure and were required to sign a confidentiality agreement. The study investigator (doctors), subjects, data managers, statisticians, and clinical research associate were all blinded to the treatment group allocation.

### Study treatment

After randomization, each patient who fulfilled with all the clinical inclusion/exclusion criteria and signed the informed consent forms received Shenfu injection (80ml Shenfu injection + 70ml 5% glucose injection) or matched placebo (150ml 5% glucose injection). The rationale behind the route of administration, timing and dosage of study medication has been previous published^20,21^. Intravenous infusion was started no more than 60 minutes before the anticipated PCI and continued for at least 30 minutes after restoration of blood flow in the culprit vessel, followed by infusion once a day and maintained for 5 days.

All patients received the standard treatments of STEMI in terms of reperfusion therapy and medical therapies according to contemporary practice guidelines and local standard of care. Other Chinese Medicine injections and oral medicines were not allowed to be used.

### Clinical follow-up

All randomized subjects underwent follow-up by telephone interviews or outpatient visit (preferred) at 30 days after randomization. The follow-up data including vital signs, laboratory tests, echocardiography, major adverse cardiovascular and cerebrovascular events (MACCE), questionnaire of quality of life, adverse event (AE), serious adverse event (SAE) and concomitant medications were collected.

### Endpoints

The primary endpoint was infarct size as a percentage of the total left ventricular (LV) mass as assessed by CMR at 5 ± 2 days after primary PCI. The secondary endpoints included microvascular obstruction and intramyocardial hemorrhage expressed as a percentage of LV mass by CMR, enzymatic infarct size by measuring the area under the curve (AUC) of creatine kinase-myocardial band (CK-MB) within 72 hours after PCI, the AUC of cardiac troponin I (cTnI/cTnT), peak value of CK-MB and peak value of cTnI/cTnT, ST segment resolution (%) immediately and 24h after PCI according to electrocardiograph, TIMI flow grade, corrected TIMI frame count (CTFC), TIMI myocardial perfusion grade (TMPG), myocardial salvage index, myocardial edema, LV end-diastolic volume, LV end-systolic volume, LV ejection fraction assessed by CMR. The incidence of MACCE (cardiovascular death, non-fatal myocardial infarction, non-fatal stroke, emergency revascularization and re-hospitalization for heart failure) and individual endpoint events at discharge and 30-day follow-up were also secondary endpoints.

### Statistical analysis

Sample size and power calculations based on our pilot data have been described previously^20, 21^. In brief, a mean infarct size in the placebo group and treatment group were 28.0% and 24.0%, respectively (responding median values are 29.1% and 24.1%, respectively), and standard deviation (SD) was 11.33^20^. We calculated that a sample size of 260 patients would provide the trial with a statistical power of at least 80% to detect an expected difference at a single-sided alpha level of 2.5%. Allowing for a drop-out rate of 20% not undergoing CMR resulted in a final sample size of 326 patients. All efficacy analyses were performed according to the intention-to-treat principle.

For continuous variables, data were expressed as mean ± SD or medians with interquartile ranges (IQR) and compared between the 2 groups using an independent-samples Student t test or the Wilcoxon rank sum test as appropriate. For categorical variables, frequencies and percentages were compared by using the Fisher exact test. Analysis of the primary endpoint and other CMR parameters was performed by using a covariance analysis model with and without adjustment for the randomization factors (center, age and TIMI flow grade pre-PCI), providing an estimated treatment effect with a 95% confidence interval of infarct size between groups. Secondary endpoints were analyzed using χ2 test or Fisher exact test for categorical data and student t test or Wilcoxon rank sum test for continuous variables, as appropriate. The following pre-specified subgroup analyses to evaluate the effect of Shenfu injection on primary endpoints were performed: age (< 65 years versus ≥ 65 years), sex, hypertension, diabetes, smoking, left ventricular ejection fraction (≤ 50% versus > 50%), time from symptom onset to reperfusion time (≤ 6 hours versus > 6 hours), Killip class (Killip class I versus Killip class > I), pre-PCI TIMI flow (TIMI 0 flow versus TIMI flow ≥1), the type of P2Y_12_ inhibitors (ticagrelor versus clopidogrel), the type of anticoagulation agents (bivalirudin versus heparin) and use of glycoprotein IIb/IIIa antagonists. All statistical analyses were performed by SAS 9.4 (SAS Institute Inc., Cary, North Carolina). All statistical tests were two-sided, and *P* < 0.05 was considered statistically significant.

### CMR Protocol and Analysis

All patients undergone ECG-gated CMR imaging with 3.0-Tesla system (Magnetom Verio, Siemens AG Healthcare, Erlangen, Germany) at 5 ± 2 days after PCI. And all CMR studies were performed blinded to treatment allocation and according to a centralized protocol. The detailed CMR protocol and analysis have been published^21^. LV function was assessed by steady-state free precession sequence cine imaging in short-axis view covering the whole LV without gap, with the parameters of repetition time (TR)=3.5, echo time (TE)=1.5 ms, 25 phases, voxel size=2.0×1.6×8 mm^3^. LV volumes, LVEF, and mass was calculated by using short-axis slices of cine images covering the whole left ventricle. Assessment of AAR was defined by a T2-weighted short-tau inversion-recovery spin echo sequence at apical, mid-ventricle, and basal level on short-axis plane, with the parameters of TR/TE 2×R-R intervals/75 ms, voxel size =2.0×1.6×8 mm^3^. AAR was considered as present if the signal intensity >2 SDs above the mean signal in remote skeletal muscle and obtained by manually tracing the hyperintense region on T2w-STIR. A hypointense area within the hyperintense region was considered as IMH. Late gadolinium enhancement images were acquired ten minutes after the intravenous administration of 0.2mmol/kg of contrast injection (Magnevist, Bayer HealthCare Pharmaceuticals Inc., Germany) by using an inversion recovery segmented 3D gradient echo sequence, with the parameters of TR=3.5, TE=1.7 ms, temporal resolution=190 ms, voxel size 1.5×1.7×10 mm3 interpolated into 0.74×0.74×5 mm3 at short-axis and 2-, 4-chamber views.

Myocardial infarction was determined as hyper-enhanced myocardium [a signal intensity >5 standard deviations (SDs) of remote normal myocardium]. MVO was determined as hypo-enhanced core within the infarcted zone. Myocardial salvage index is calculated as: [(AAR – infarct size)/AAR]x100%. The extent of the infarction, MVO, IMH and AAR was expressed as a percentage of LV myocardial mass (%LV). All CMR imaging was analyzed offline by blinded expert observers using CVI42 Version 5.13 (Cardiovascular Imaging, Calgary, Alberta, Canada).

## Results

### Patients Characteristics

The study flowchart is shown in Figure 1. A total of 351 patients planned to undergoing primary PCI for acute anterior STEMI were screened and 295 patients were randomized (144 to the Shenfu injection group; 151 to the placebo group). The patients with a missing primary endpoint (infarct size measured by CMR) comprised 10 patients in the Shenfu injection and 12 patients in the placebo group. Thus, 273 patients (134 patients in the Shenfu injection group and 139 patients in the placebo group) underwent CMR imaging and had the available data for the primary analysis.

**Figure 1:**
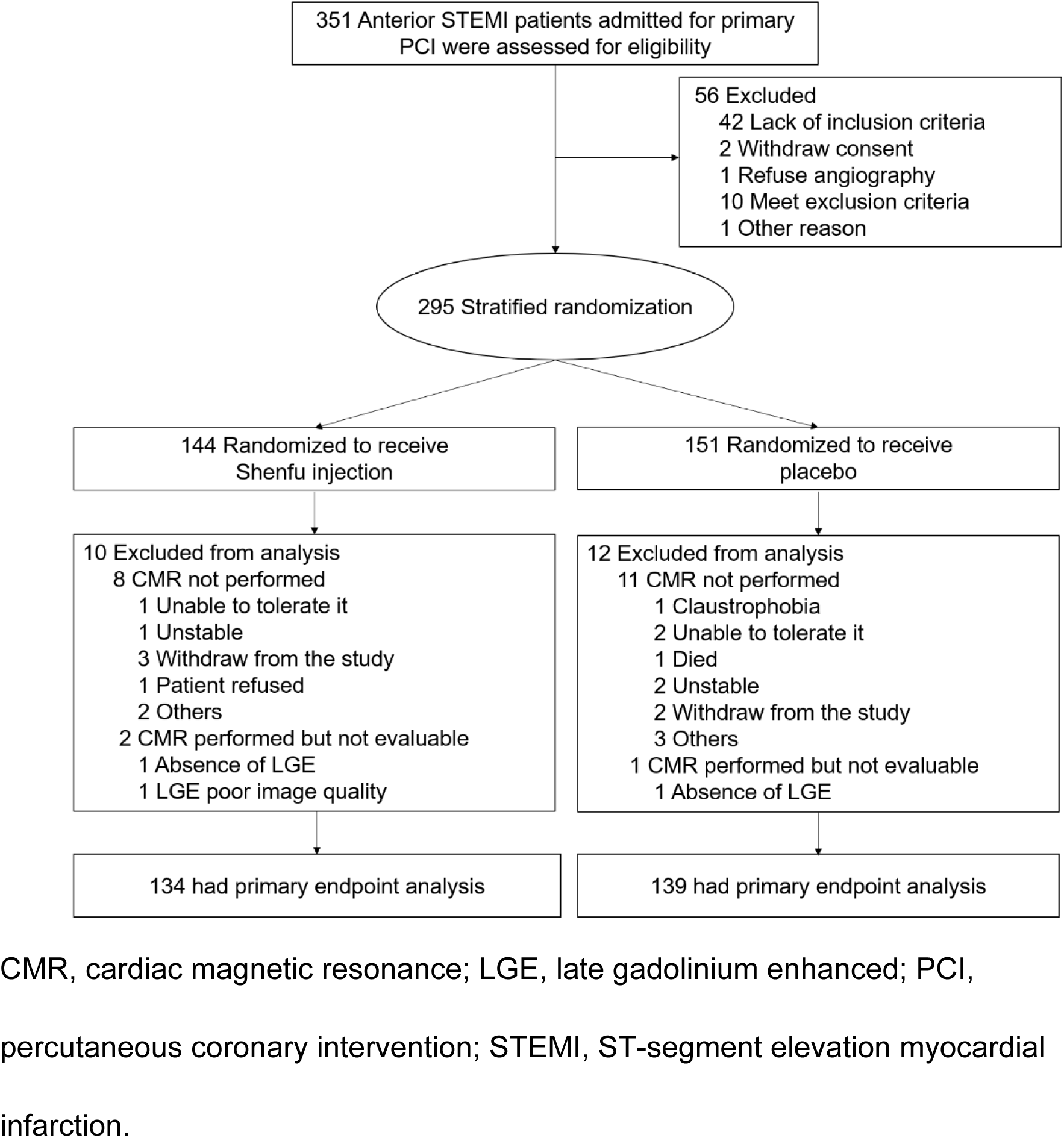
Flowchart of the RESTORE trial.

The baseline characteristics of patients are shown in Table 1. The median age of the study population was 56.5 (IQR: 48.0-65.0) years, and 83.4% were male. Except for age, there were no major significances at baseline between the two groups.

**Table 1.**
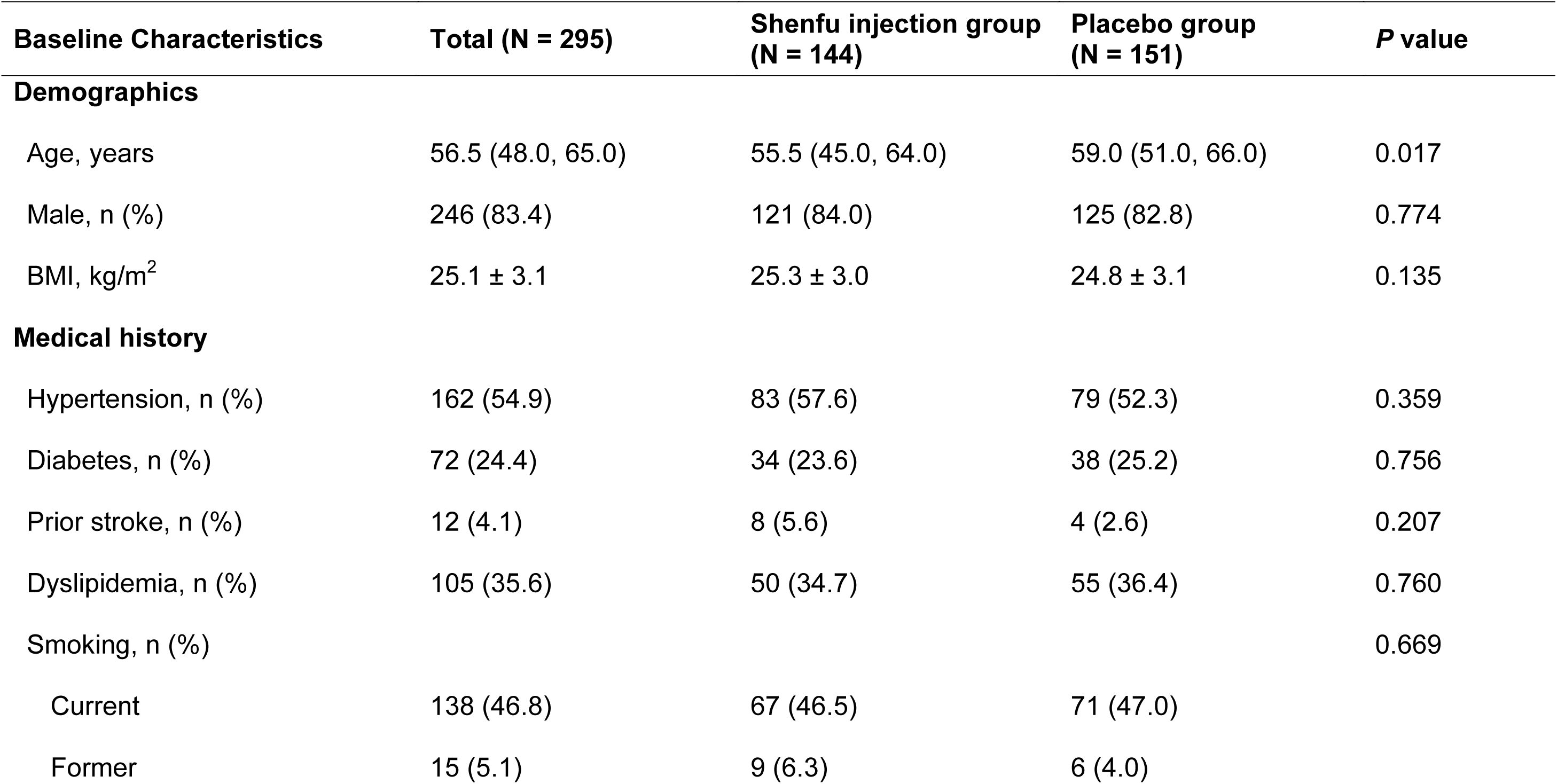
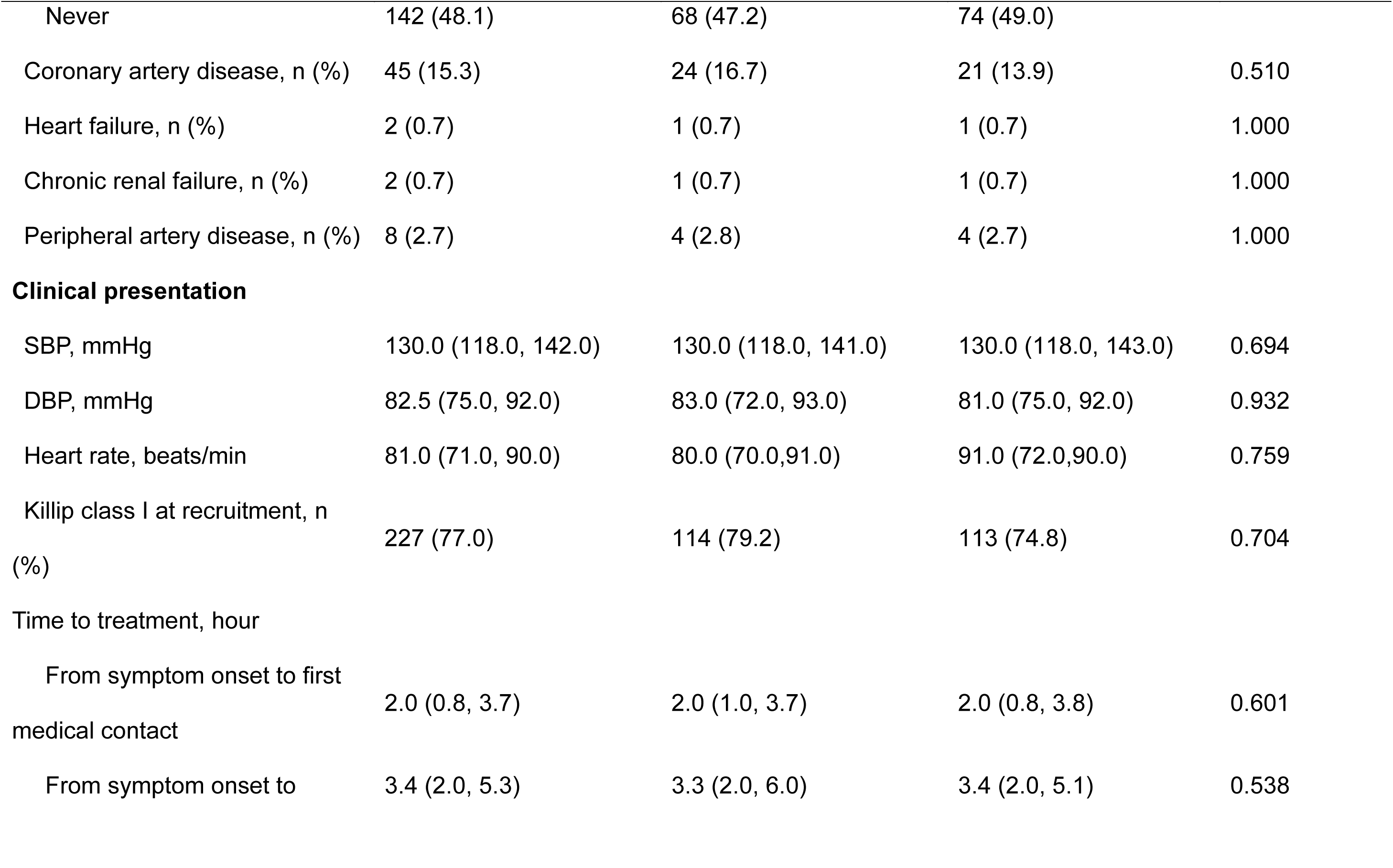
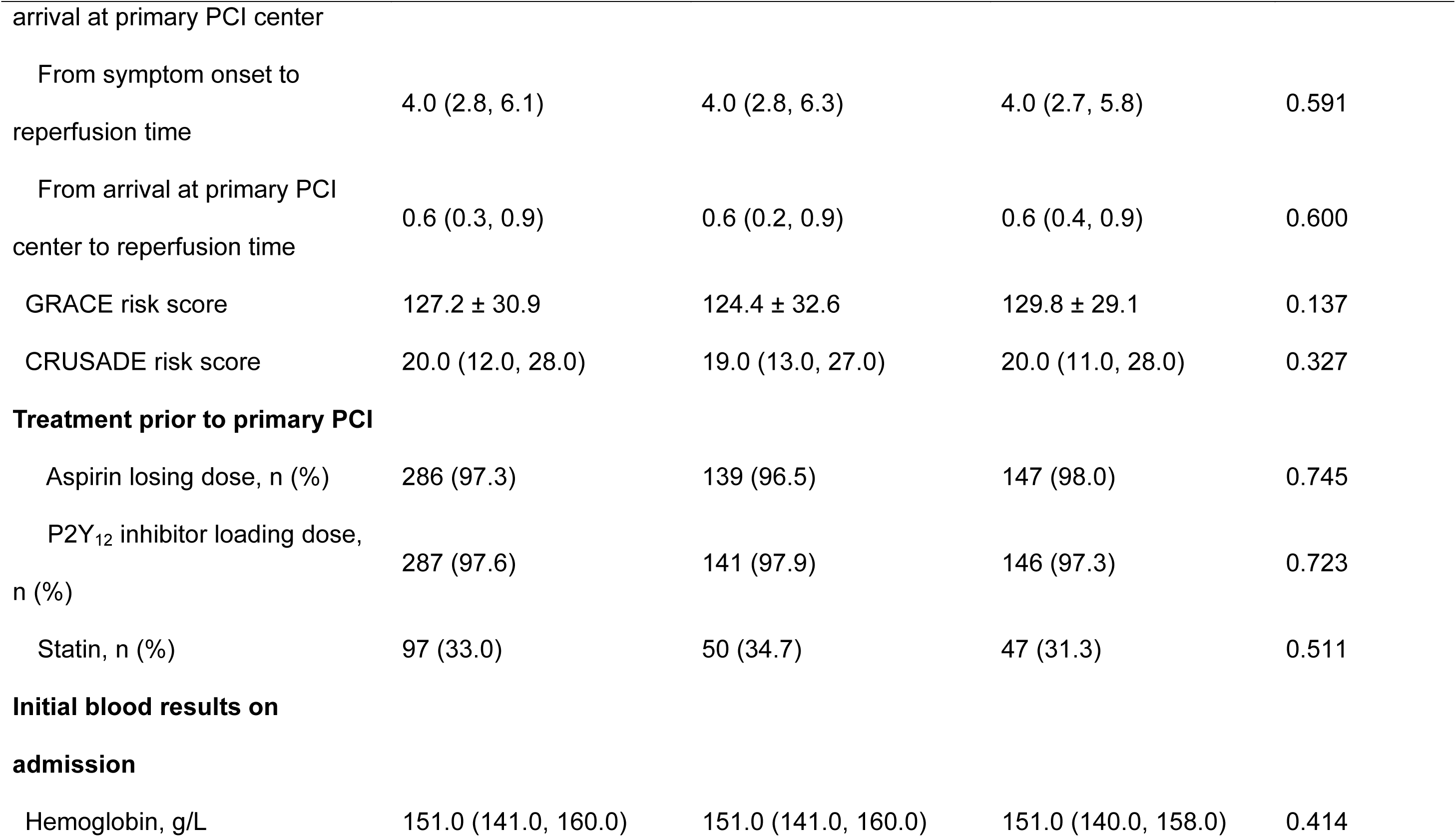
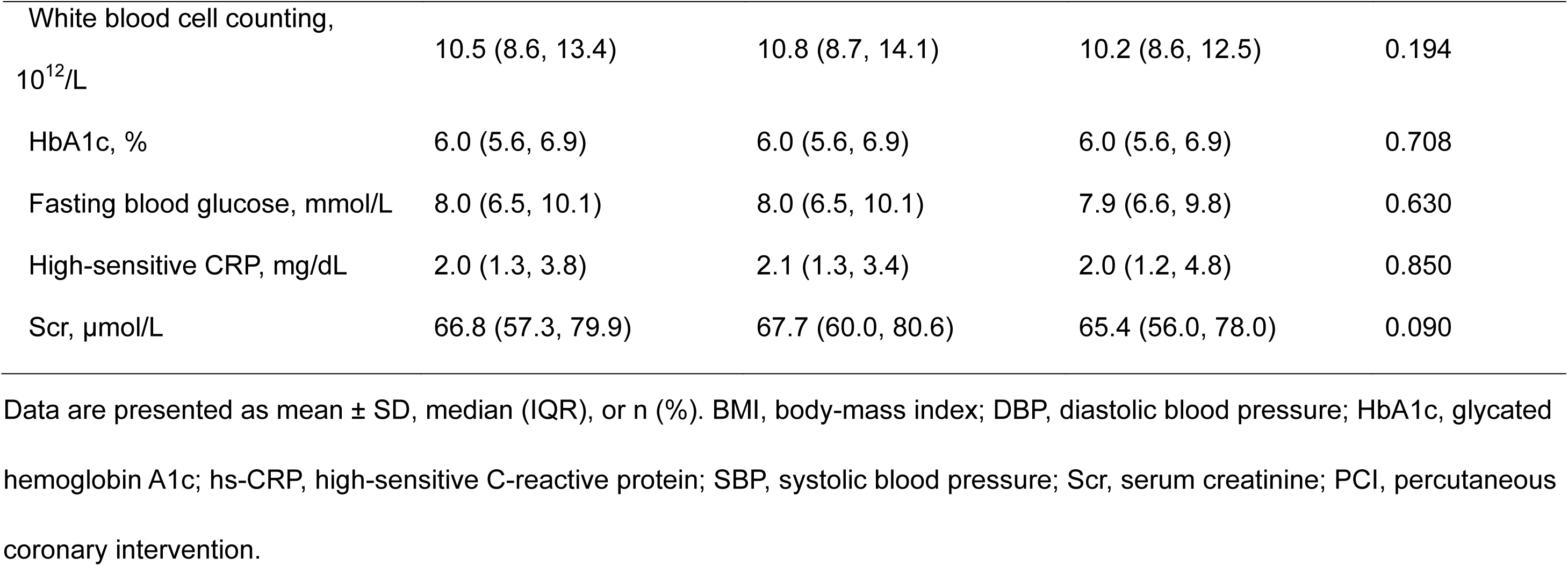
Baseline Clinical Characteristics.

Among included patients, 54.9% had hypertension, 24.4% had diabetes, and 15.3% presented with known coronary artery disease. The two groups had a similar hemodynamic status, in terms of blood pressure, heart rate and Killip class. Baseline risk score, pharmacological treatment prior to PCI, different time intervals concerning treatment, and blood results on admission were also well balanced between groups.

Procedural and angiographic data are shown in Table 2. Distribution of infarct-related artery (LAD) stenosis location and number of diseased arteries were similar between groups. There was a trend towards a higher incidence of pre-PCI TIMI 0 in the Shenfu injection group compared with that in the placebo group. Stent implantation was performed in 91.9% of patients, and drug-eluting stent was used in all patients with stent implantation. Interventional and pharmacological treatment at discharge were comparable between the two groups.

**Table 2.**
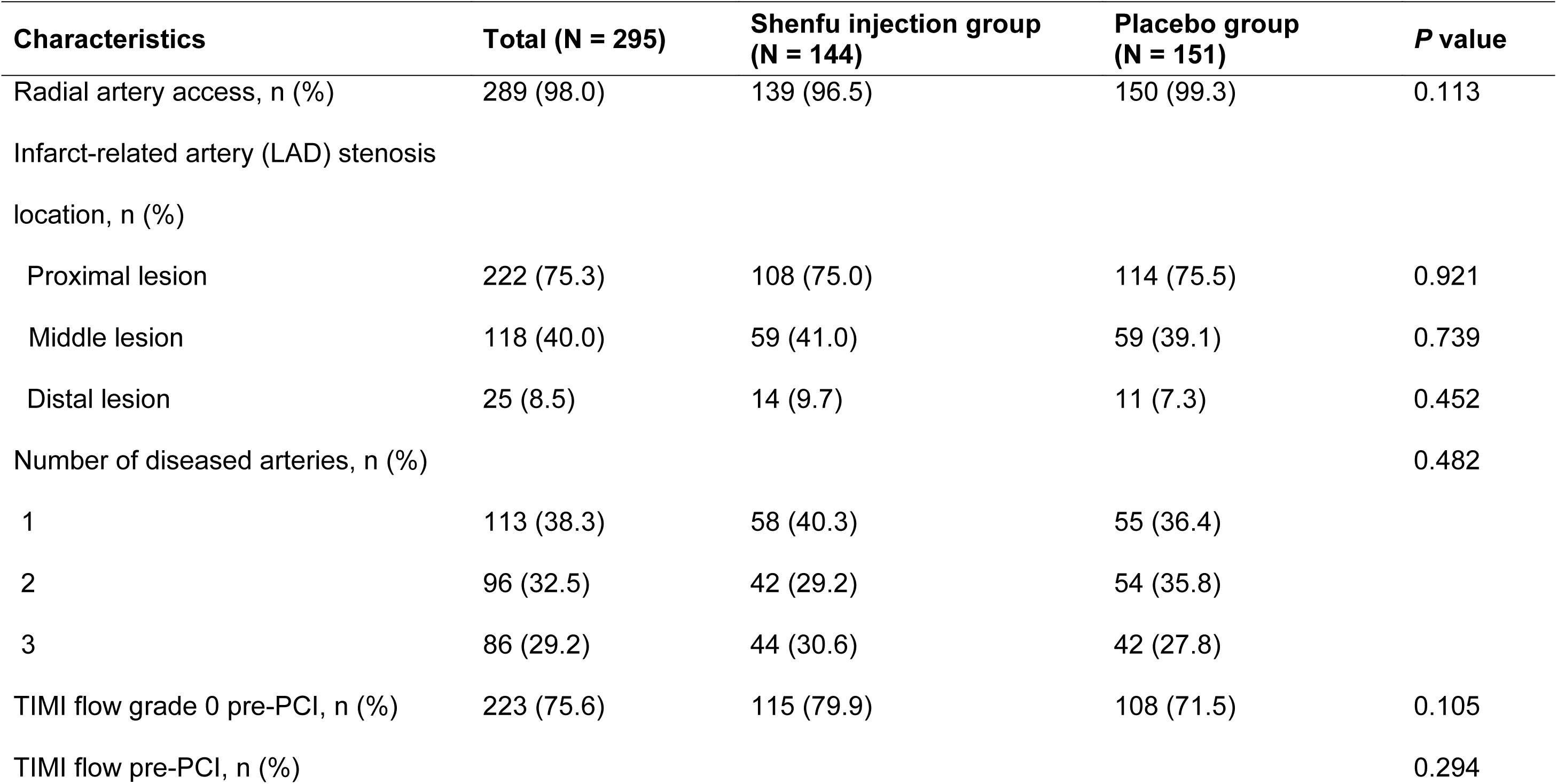
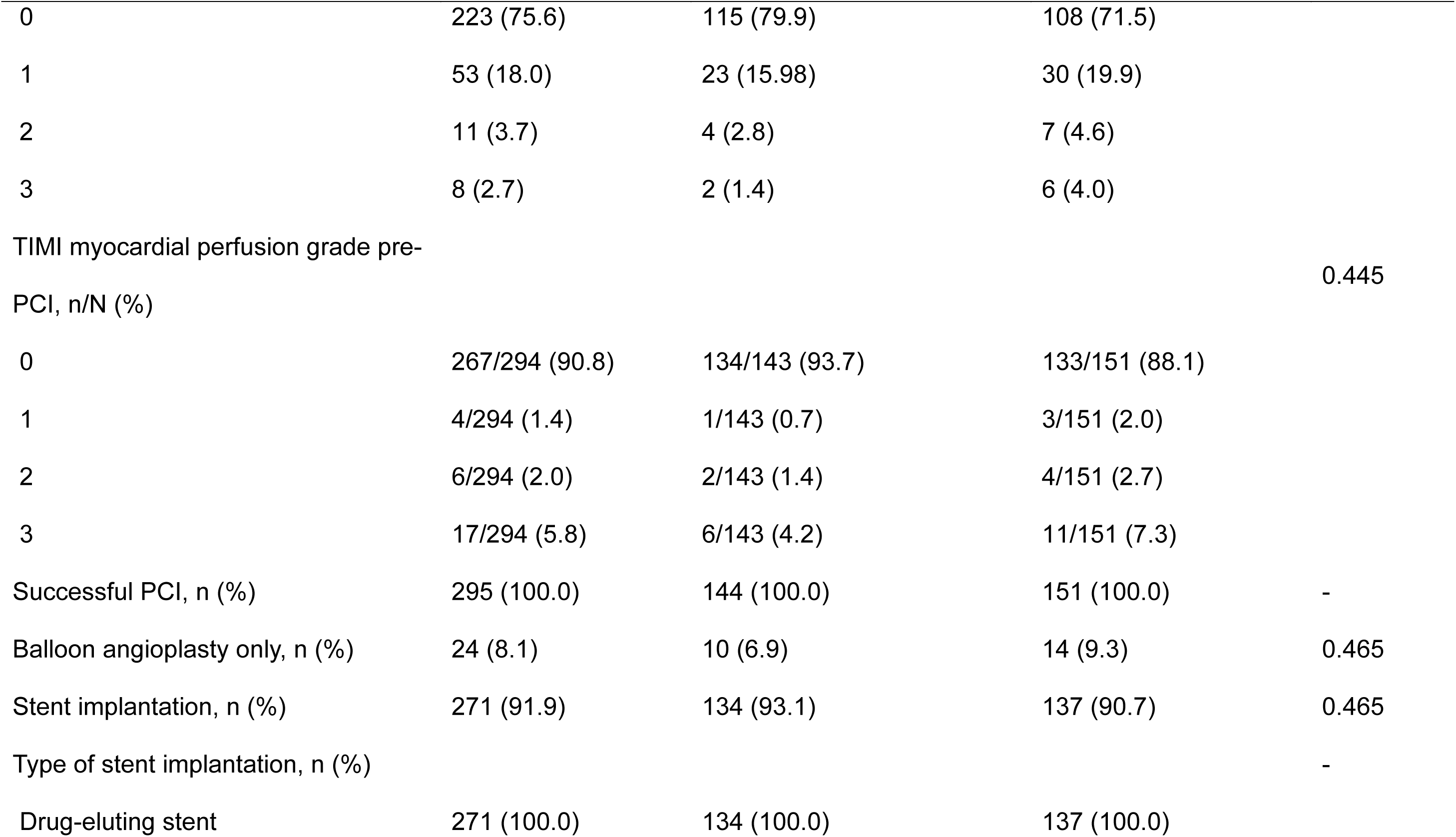
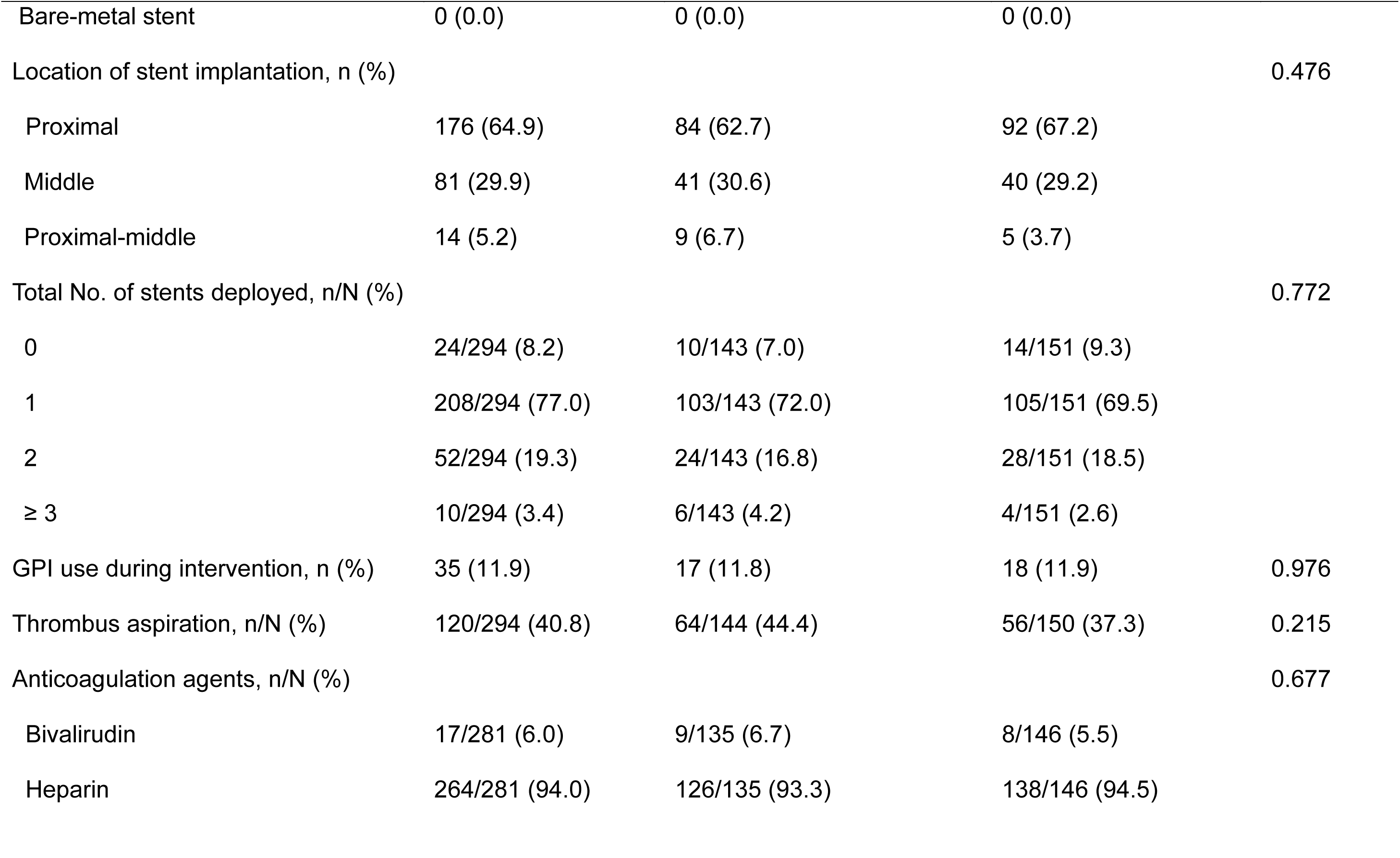
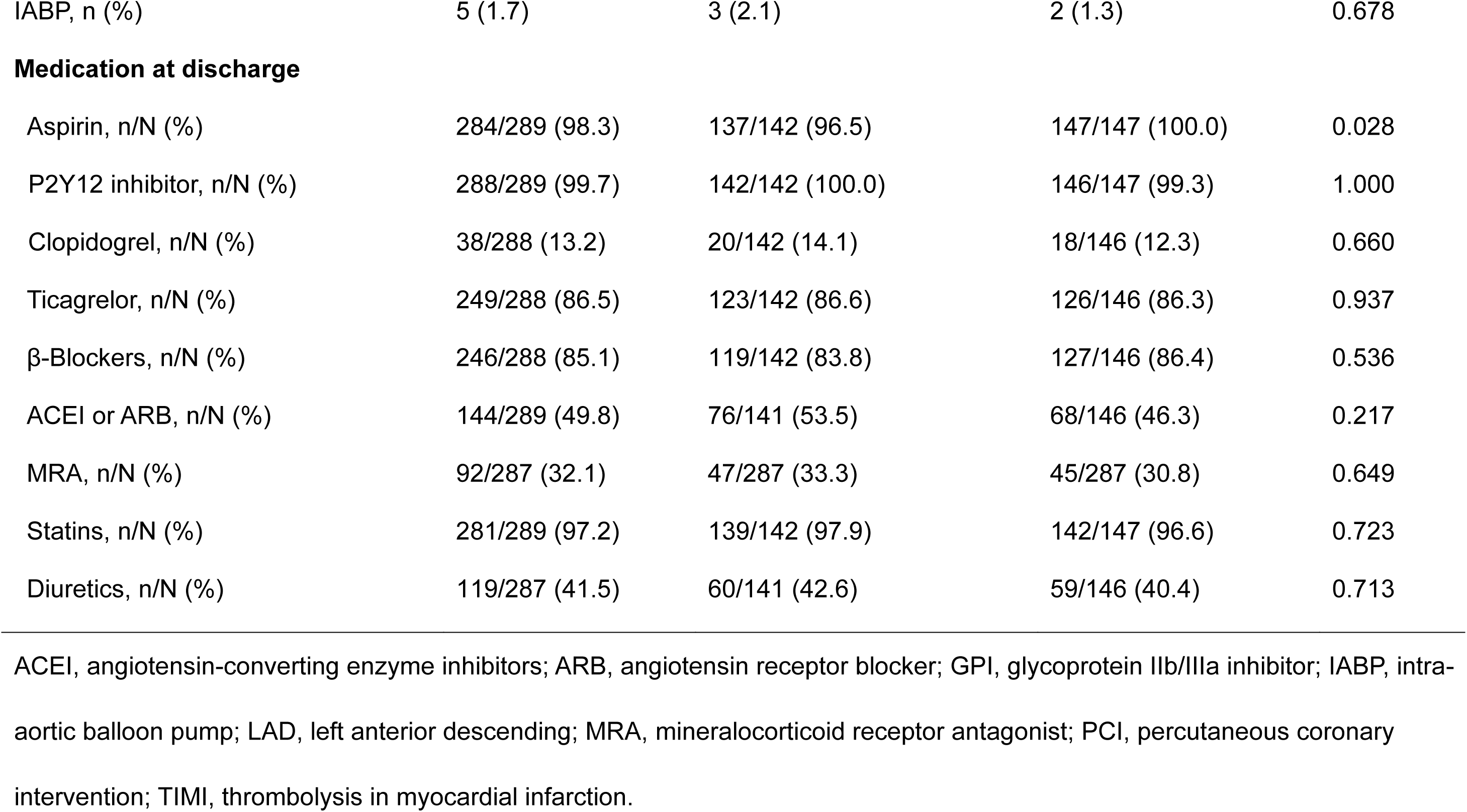
Procedural Characteristics and Medication at Discharge.

| **Characteristics** | **Total (N = 295)** | **Shenfu injection group**<br>**(N = 144)** | **Placebo group**<br>**(N = 151)** | ***P* value** |
| --- | --- | --- | --- | --- |
| Radial artery access, n (%) | 289 (98.0) | 139 (96.5) | 150 (99.3) | 0.113 |
| Infarct-related artery (LAD) stenosis<br>location, n (%) |  |  |  |  |
| Proximal lesion | 222 (75.3) | 108 (75.0) | 114 (75.5) | 0.921 |
| Middle lesion | 118 (40.0) | 59 (41.0) | 59 (39.1) | 0.739 |
| Distal lesion | 25 (8.5) | 14 (9.7) | 11 (7.3) | 0.452 |
| Number of diseased arteries, n (%) |  |  |  | 0.482 |
| 1 | 113 (38.3) | 58 (40.3) | 55 (36.4) |  |
| 2 | 96 (32.5) | 42 (29.2) | 54 (35.8) |  |
| 3 | 86 (29.2) | 44 (30.6) | 42 (27.8) |  |
| TIMI flow grade 0 pre-PCI, n (%) | 223 (75.6) | 115 (79.9) | 108 (71.5) | 0.105 |
| TIMI flow pre-PCI, n (%) |  |  |  | 0.294 |
| 0 | 223 (75.6) | 115 (79.9) | 108 (71.5) |  |
| 1 | 53 (18.0) | 23 (15.98) | 30 (19.9) |  |
| 2 | 11 (3.7) | 4 (2.8) | 7 (4.6) |  |
| 3 | 8 (2.7) | 2 (1.4) | 6 (4.0) |  |
| TIMI myocardial perfusion grade pre-PCI, n/N (%) |  |  |  | 0.445 |
| 0 | 267/294 (90.8) | 134/143 (93.7) | 133/151 (88.1) |  |
| 1 | 4/294 (1.4) | 1/143 (0.7) | 3/151 (2.0) |  |
| 2 | 6/294 (2.0) | 2/143 (1.4) | 4/151 (2.7) |  |
| 3 | 17/294 (5.8) | 6/143 (4.2) | 11/151 (7.3) |  |
| Successful PCI, n (%) | 295 (100.0) | 144 (100.0) | 151 (100.0) | - |
| Balloon angioplasty only, n (%) | 24 (8.1) | 10 (6.9) | 14 (9.3) | 0.465 |
| Stent implantation, n (%) | 271 (91.9) | 134 (93.1) | 137 (90.7) | 0.465 |
| Type of stent implantation, n (%) |  |  |  | - |
| Drug-eluting stent | 271 (100.0) | 134 (100.0) | 137 (100.0) |  |
| Bare-metal stent | 0 (0.0) | 0 (0.0) | 0 (0.0) |  |
| Location of stent implantation, n (%) |  |  |  | 0.476 |
| Proximal | 176 (64.9) | 84 (62.7) | 92 (67.2) |  |
| Middle | 81 (29.9) | 41 (30.6) | 40 (29.2) |  |
| Proximal-middle | 14 (5.2) | 9 (6.7) | 5 (3.7) |  |
| Total No. of stents deployed, n/N (%) |  |  |  | 0.772 |
| 0 | 24/294 (8.2) | 10/143 (7.0) | 14/151 (9.3) |  |
| 1 | 208/294 (77.0) | 103/143 (72.0) | 105/151 (69.5) |  |
| 2 | 52/294 (19.3) | 24/143 (16.8) | 28/151 (18.5) |  |
| ≥ 3 | 10/294 (3.4) | 6/143 (4.2) | 4/151 (2.6) |  |
| GPI use during intervention, n (%) | 35 (11.9) | 17 (11.8) | 18 (11.9) | 0.976 |
| Thrombus aspiration, n/N (%) | 120/294 (40.8) | 64/144 (44.4) | 56/150 (37.3) | 0.215 |
| Anticoagulation agents, n/N (%) |  |  |  | 0.677 |
| Bivalirudin | 17/281 (6.0) | 9/135 (6.7) | 8/146 (5.5) |  |
| Heparin | 264/281 (94.0) | 126/135 (93.3) | 138/146 (94.5) |  |
| IABP, n (%) | 5 (1.7) | 3 (2.1) | 2 (1.3) | 0.678 |
| **Medication at discharge** |  |  |  |  |
| Aspirin, n/N (%) | 284/289 (98.3) | 137/142 (96.5) | 147/147 (100.0) | 0.028 |
| P2Y12 inhibitor, n/N (%) | 288/289 (99.7) | 142/142 (100.0) | 146/147 (99.3) | 1.000 |
| Clopidogrel, n/N (%) | 38/288 (13.2) | 20/142 (14.1) | 18/146 (12.3) | 0.660 |
| Ticagrelor, n/N (%) | 249/288 (86.5) | 123/142 (86.6) | 126/146 (86.3) | 0.937 |
| β-Blockers, n/N (%) | 246/288 (85.1) | 119/142 (83.8) | 127/146 (86.4) | 0.536 |
| ACEI or ARB, n/N (%) | 144/289 (49.8) | 76/141 (53.5) | 68/146 (46.3) | 0.217 |
| MRA, n/N (%) | 92/287 (32.1) | 47/287 (33.3) | 45/287 (30.8) | 0.649 |
| Statins, n/N (%) | 281/289 (97.2) | 139/142 (97.9) | 142/147 (96.6) | 0.723 |
| Diuretics, n/N (%) | 119/287 (41.5) | 60/141 (42.6) | 59/146 (40.4) | 0.713 |

ACEI, angiotensin-converting enzyme inhibitors; ARB, angiotensin receptor blocker; GPI, glycoprotein IIb/IIIa inhibitor; IABP, intra-aortic balloon pump; LAD, left anterior descending; MRA, mineralocorticoid receptor antagonist; PCI, percutaneous coronary intervention; TIMI, thrombolysis in myocardial infarction.

The baseline characteristics, procedural and angiographic data of patients who completed CMR for primary analysis are shown in Table S2 and Table S3.

### CMR assessment

Of 295 patients, 276 (93.6%) underwent CMR at a median of 5 days (IQR: 4-5). For the primary endpoint (CMR-defined infarct size), 134 of 144 patients in the Shenfu injection group and 139 of 151 patients in the placebo group had an accurate endpoint assessment, respectively.

### Primary endpoint

The primary endpoint of infarct size expressed as a percentage of LV mass is shown in Table 3, Figure 2, Central Illustration and Supplemental Table 4. The unadjusted infarct size revealed by CMR was not significantly reduced in the Shenfu injection group compared with the placebo group (37.4% versus 37.5%; mean difference, - 0.04%; 95% *CI*: −3.45% to 3.37%; *P* = 0.982). On covariance analysis model with adjustment for the randomization factors, the adjusted infarct size between groups remained no significant difference (30.9% versus 31.2%; mean difference, −0.26%; 95%*CI*: −3.61% to 3.08%; *P* = 0.877).

**Figure 2:**
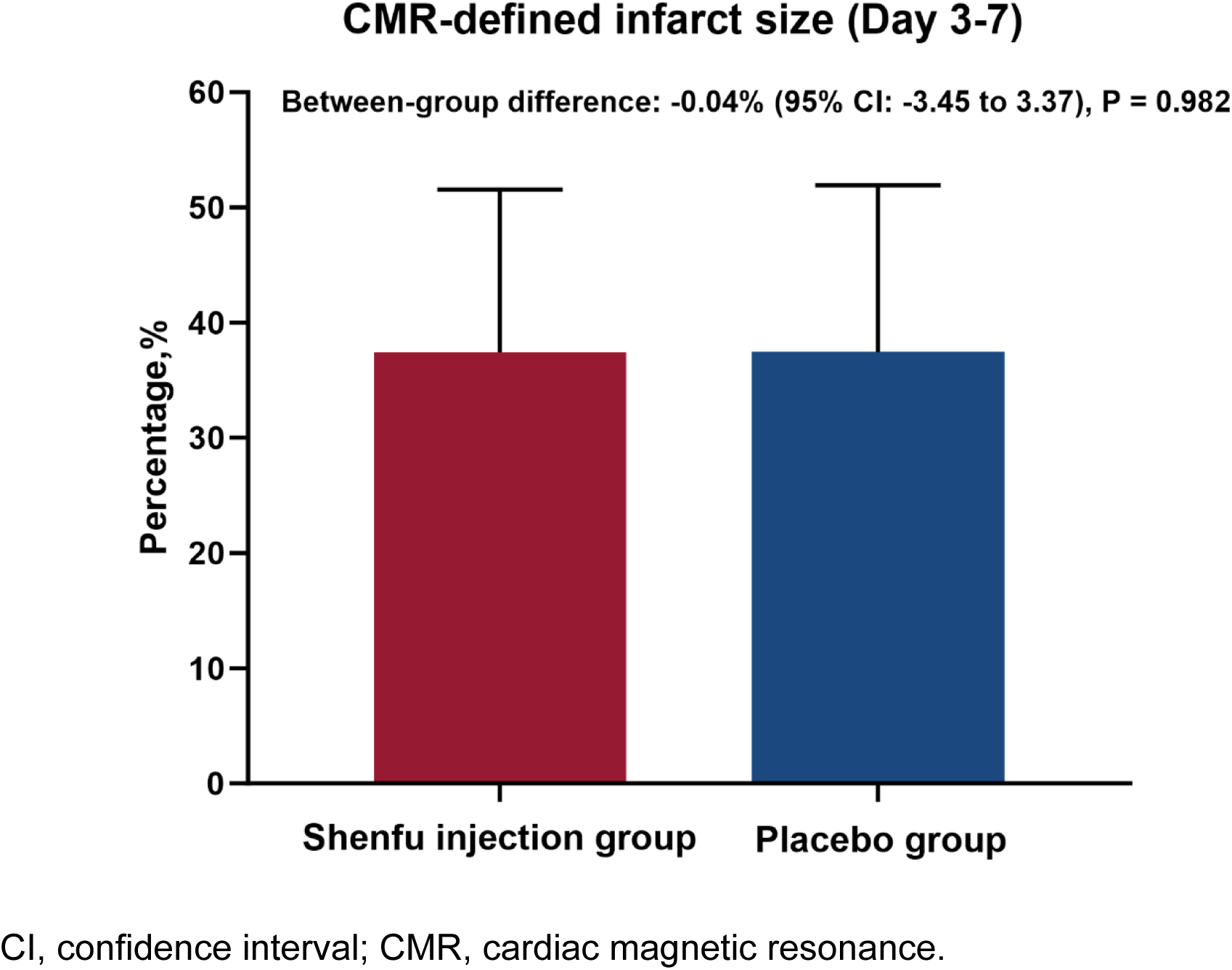
Primary endpoint analyses.

**Table 3.** Prespecified Primary and Secondary Endpoints.

| **Outcomes** | **Shenfu injection group**<br>**(N = 134)** | **Placebo group**<br>**(N = 139)** | **Between-group**<br>**difference (95% CI)** | **P value** |
| --- | --- | --- | --- | --- |
| **Primary endpoint** |  |  |  |  |
| Infarct size at 5 ± 2 days, % LV mass | 37.4 ± 14.1 | 37.5 ± 14.5 | -0.04(-3.45,3.37) | 0.982 |
| **Secondary endpoints** |  |  |  |  |
| MVO at 5 ± 2 days, % LV mass | 1.7 (0.0, 4.6) | 0.7 (0.0, 4.6) | 0.39 (-0.60, 1.39) | 0.118 |
| IMH at 5 ± 2 days, % LV mass | 0.1 (0.0, 1.4) | 0.0 (0.0, 0.9) | 0.18 (-0.21, 0.58) | 0.153 |
| MSI at 5 ± 2 days, % | 36.7 ± 18.4 | 38.0 ± 17.7 | -1.39 (-5.72, 2.95) | 0.529 |
| AAR at 5 ± 2 days, % LV mass | 59.9 (50.6, 68.0) | 60.2 (53.0, 67.1) | -0.75 (-4.15, 2.65) | 0.654 |
| LVEF at 5 ± 2 days, % | 47.5 ± 9.8 | 47.9 ± 10.5 | -0.40 (-2.82, 2.02) | 0.747 |
| LVEDV at 5 ± 2 days, mL | 129.8 ± 29.7 | 131.7 ± 31.3 | -1.84 (-9.13, 5.44) | 0.619 |
| LVESV at 5 ± 2 days, mL | 68.8 ± 22.9 | 70.1 ± 27.0 | -1.23 (-7.22, 4.75) | 0.800 |
| TIMI flow 3 post-PCI, n (%) | 127 (94.8) | 135 (97.1) | - | 0.324 |
| TIMI myocardial perfusion grade 3 post-PCI, n/N (%) | 122/133 (91.7) | 128/139 (92.1) | - | 0.914 |
| Corrected TIMI frame count post-PCI | 11 (8, 15) | 11.5 (8, 16) | -0.22(-1.93, 1.49) | 0.913 |
| Peak value of CK-MB, ng/mL | 217.3 (86.4, 293.0) | 193.2 (80.0, 290.7) | - | 0.445 |
| Peak value of cTnT, ng/mL | 5.7 (3.7, 10.0) | 8.73 (4.5, 10.0) | - | 0.884 |
| CK-MB, AUC | 774.8 (361.0, 1155.0) | 735.1 (334.2, 1021.8) | - | 0.337 |
| cTnT, AUC | 35.0 (20.2, 67.1) | 38.7 (16.0, 54.6) | - | 0.917 |
| Complete ST-segment resolution<br>immediately post-PCI, n/N (%) | 29/114 (25.4) | 29/111 (26.1) | - | 0.906 |
| Complete ST-segment resolution 24h<br>post-PCI | 45/109 (41.3) | 46/113 (40.7) | - | 0.930 |
| Slow flow during procedure, n (%) | 4 (3.0) | 6 (4.3) | - | 0.750 |
| No-flow during procedure, n (%) | 0 (0.0) | 1 (0.7) | - | 1.000 |
| Malignant arrhythmia during PCI, n (%) | 0 (0.0) | 0 (0.0) | - | - |
| Hs-CRP, mg/dL |  |  |  |  |
| Hs-CRP 24 hours after PCI | 10.9 (5.0, 20.4) | 11.6 (5.6, 20.0) | -154.06 (-445.82, 137.71) | 0.709 |
| Hs-CRP 72 hours after PCI | 14.4 (8.1, 32.8) | 17.3 (7.7, 36.1) | -4.04 (-11.95, 3.87) | 0.465 |
| Hs-CRP 5 days after PCI | 9.7 (5.4, 23.2) | 11.0 (5.2, 24.6) | -2.15 (-7.84, 3.53) | 0.944 |
| BNP, pg/mL |  |  |  |  |
| BNP 24 hours after PCI | 363.0 (196.0, 740.0) | 347.5 (179.8, 668.0) | 55.87 (-249.42, 361.16) | 0.738 |
| BNP 72 hours after PCI | 200.0 (98.0, 515.0) | 194.0 (111.0, 623.0) | -64.47 (-466.67, 337.74) | 0.775 |
| BNP 5 days after PCI | 211.0 (114.8, 441.0) | 198.0 (109.8, 409.0) | -3.59 (-227.72, 220.55) | 0.768 |
| NT-proBNP, pg/mL |  |  |  |  |
| NT-proBNP 24 hours after PCI | 1420.0 (772.0, 2604.0) | 1214.0 (848.0, 1849.0) | 530.30 (-277.33, 1337.94) | 0.572 |
| NT-proBNP 72 hours after PCI | 743.0 (267.0, 1378.0) | 950.0 (594.0, 1324.0) | -285.50 (-1205.71, 634.72) | 0.345 |
| NT-proBNP 5 days after PCI | 633.0 (226.0, 1120.0) | 645.5 (419.0, 1079.0) | -82.30 (-710.60, 546.01) | 0.521 |
| MIDAS score 6h post-PCI | 59 (46, 98) | 61 (47, 98) | -0.74 (-7.81, 6.34) | 0.780 |
| MIDAS score 30d post-PCI | 53 (40, 74) | 54 (71, 78) | -0.69 (-5.77, 4.39) | 0.928 |

AAR, area at risk; AUC, area under curve; BNP, brain natriuretic peptide; CK-MB, creatine kinase-myocardial band; cTnI, cardiac troponin I; cTnT, cardiac troponin T; hs-CRP, high-sensitive C-reactive protein; IMH, intra-myocardial hemorrhage; LV, left ventricular; LVEDV, left ventricular end diastolic volume; LVEF, left ventricular ejection fraction; LVESV, left ventricular end systolic volume; MIDAS, myocardial infarction dimensional assessment scale; MVO, microvascular obstruction; MSI, myocardial salvage index; NT-proBNP, N-terminal pro brain natriuretic peptide; PCI, percutaneous coronary intervention; TIMI, thrombolysis in myocardial infarction.

There were no significant differences across the pre-specified subgroups for the primary endpoint (Figure 3). Notably, in the subgroup of patients with age ≥ 65 years, infarct size was numerically but not significantly decreased in the Shenfu injection group (36.2% versus 40.2%, or a 3.99 reduction [95%*CI*: −10.27% to 2.29%], *P* = 0.209).

**Figure 3:**
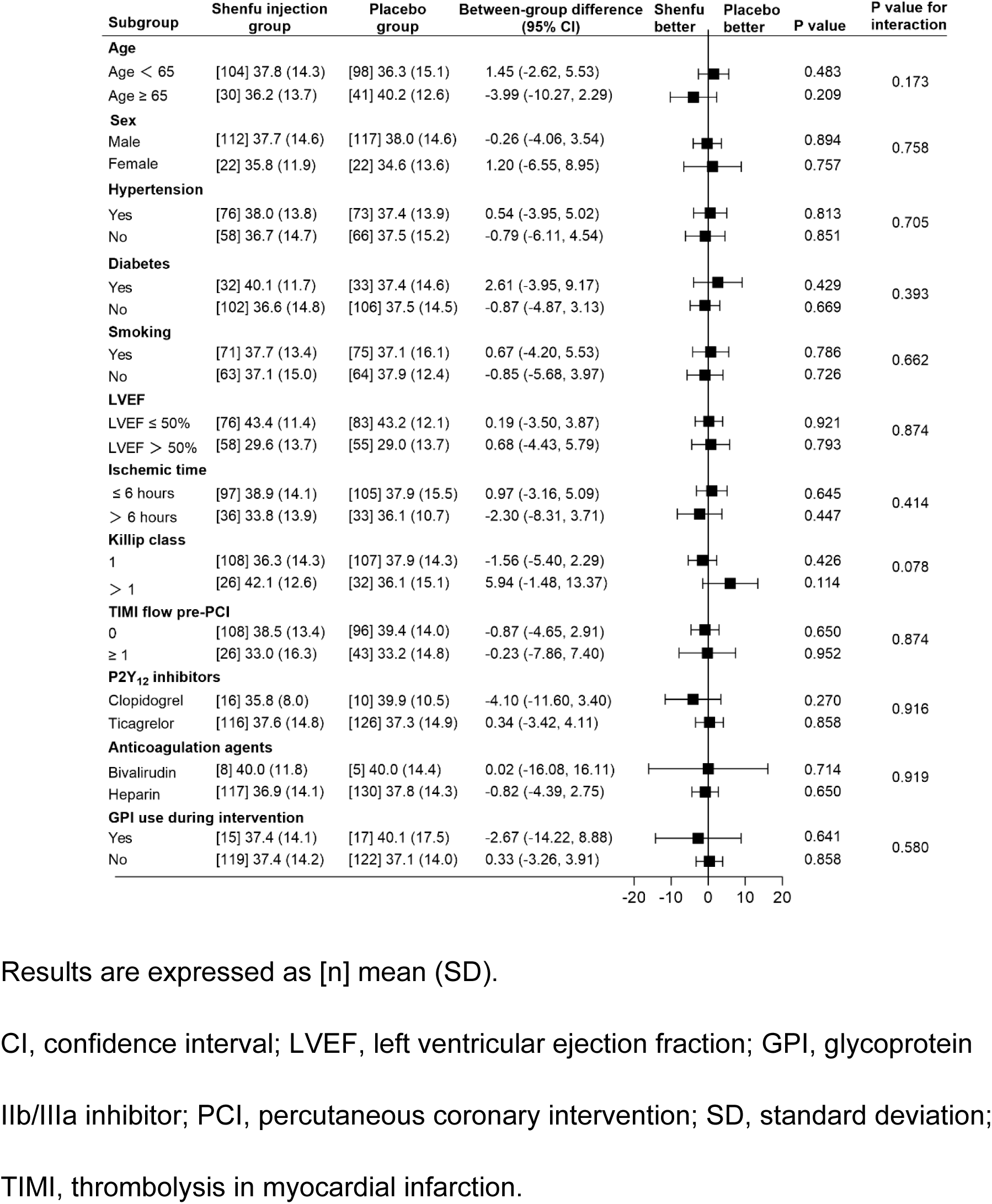
Forest Plot for the Effect of Shenfu Injection on Infarct Size in Pre-specified Subgroups.

### Secondary endpoints

There were no significant differences in the pre-specified secondary endpoints defined by CMR, in terms of microvascular obstruction, intramyocardial hemorrhage, myocardial salvage index, myocardial edema, LV end-diastolic volume, LV end-systolic volume, and LV ejection fraction between the Shenfu injection and placebo groups with and without adjustment, as shown in Table 3 and Table S4.

There were no significant differences by study treatment in the incidence of the coronary-related secondary endpoints, including TIMI flow 3 post-PCI (94.8% versus 97.1%, *P* = 0.324), TMPG 3 post-PCI (91.7% versus 92.1%, *P* = 0.914), no-reflow (0.0% versus 0.7%, *P* = 1.000), and slow-flow (3.0% versus 4.3%, *P* = 0.750). Corrected TIMI frame count post-PCI, as a continue assessment for epicardial reperfusion, showed no significant differences between the two treatment arms. Infarct size by the AUC of CK-MB or cTnT over the initial 72 h post-PCI did not differ between groups (Figure 4), as well as the peak value of CKMB and cTnT. Complete ST-segment resolution immediately or at 24h post-PCI was also similar between groups. High-sensitive C-reactive protein levels and BNP or NT-proBNP were not significantly lower in the Shenfu injection group than in the placebo group at 24 hours, 72 hours and 5 days. Health-related quality of life scores (MIDAS score) was not significantly different between groups at 6h or 30 days.

**Figure 4:**
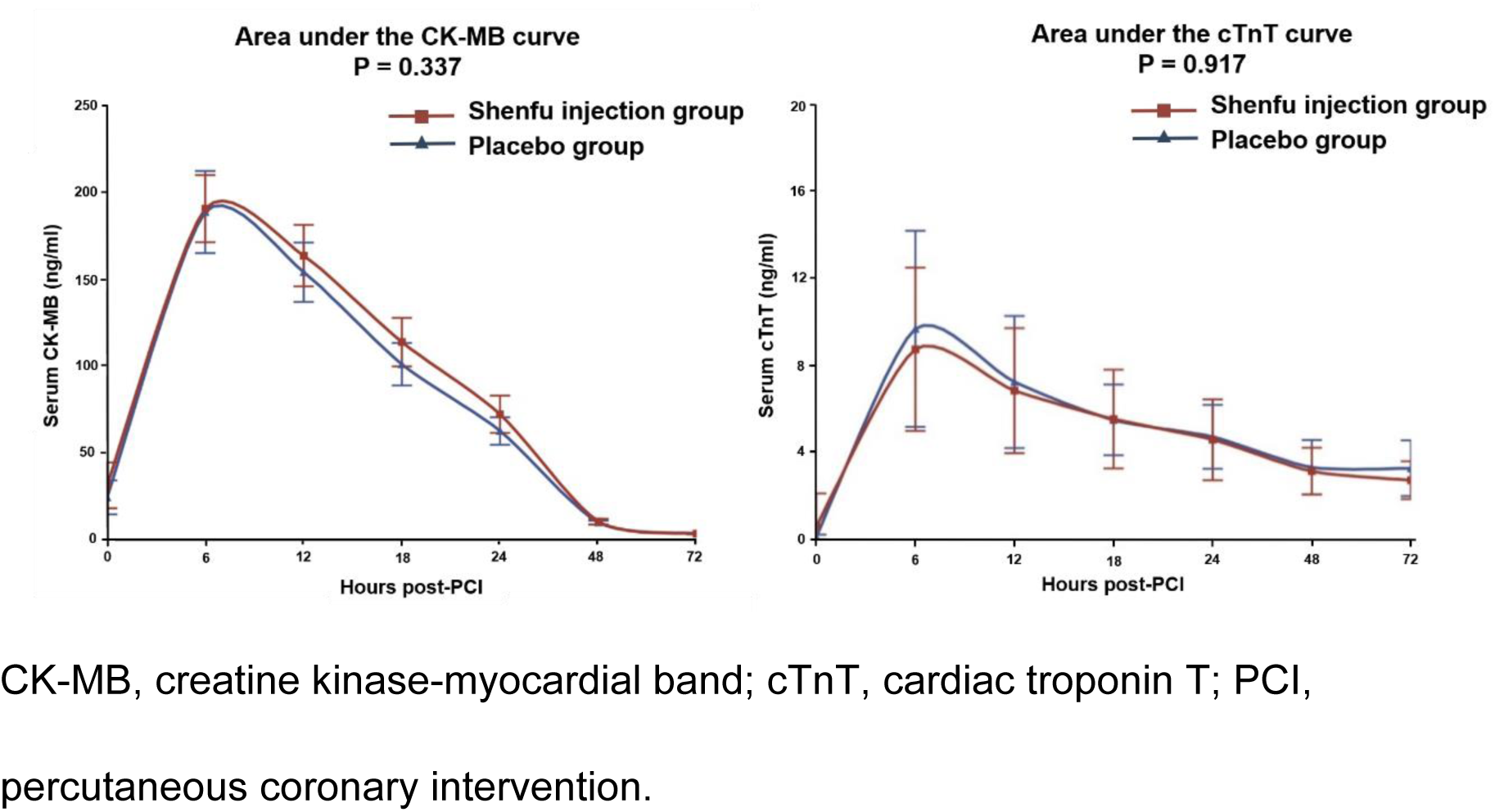
Infarct Size by the Area Under the Curve of CK-MB and cTnT.

### Per-protocol analysis

Following per-protocol analysis, there remained no reduction in infarct size in Shenfu injection group compared with placebo group in unadjusted or adjusted analyses, as well as other CMR parameters, as shown in Table S5, Table S6 and Figure S1.

Similarly, there were no significant differences between groups in terms of infarct size by the AUC of CK-MB or cTnT over the initial 72h post-PCI (Figure S2).

### Clinical outcomes at discharge and at 30-day, severe adverse events, and adverse events

The clinical outcomes are described in Table 4. No significant difference was found for the rate of MACCE at 30 days. At discharge, MACCE occurred in 2 patient (1.3%) in the placebo group, and no MACCE occurred in the Shenfu injection group.

**Table 4.** Main Adverse Cardiovascular and Cerebrovascular at Discharge and 30 Days after PCI.

| **Main adverse cardiovascular and cerebrovascular events** | **Shenfu injection group (N = 144)** | **Placebo group (N = 151)** | ***P* value** |
| --- | --- | --- | --- |
| MACCE at discharge, n (%) | 0 (0.0) | 2 (1.3) | 0.499 |
| Cardiovascular death, n (%) | 0 (0.0) | 1 (0.7) | 1.000 |
| Non-fatal myocardial infarction, n (%) | 0 (0.0) | 0 (0.0) | - |
| Non-fatal stroke, n (%) | 0 (0.0) | 0 (0.0) | - |
| Emergency revascularization, n (%) | 0 (0.0) | 1 (0.7) | 1.000 |
| MACCE at 30 days after PCI, n (%) | 2 (1.4) | 4 (2.6) | 0.685 |
| Cardiovascular death, n (%) | 1 (0.7) | 2 (1.3) | 1.000 |
| Non-fatal myocardial infarction, n (%) | 0 (0.0) | 0 (0.0) | - |
| Non-fatal stroke, n (%) | 1 (0.7) | 0 (0.0) | 0.488 |
| Emergency revascularization, n (%) | 0 (0.0) | 1 (0.7) | 1.000 |
| Re-hospitalization for heart failure, n (%) | 0 (0.0) | 1 (0.7) | 1.000 |

MACCE, main adverse cardiovascular and cerebrovascular events; PCI, percutaneous coronary intervention.

By 30 days, 1 patient (0.7%) in the Shenfu injection group and 3 patient (2.0%) in the placebo group had died. Three deaths were secondary to cardiovascular causes (1 probable subacute stent thrombosis in the Shenfu injection group, and 1 ventricular fibrillation and 1 cardiac rupture in the placebo group), another death was attributable to infectious pneumonia in the placebo group. The rate of MACCE at 30 days was not significantly different between the two groups: 2 patient (1.4%) in the Shenfu injection group and 4 patients (2.6%) in the placebo group, respectively (*P* = 0.685).

There were 9 patients (6.25%) with severe adverse events in the Shenfu injection group and 11 patients (7.28%) in the placebo group at 30-day follow-up, with no significant difference (*P* = 0.819, Table 5). Adverse events classified according to System Organ Class are shown in Table S7. Rates of adverse events were similar between the two groups (61.1% versus 63.6%, *P* = 0.719).

**Table 5.** Severe Adverse Events.

| **Events** | **Shenfu injection group**<br>**(N = 144)** | **Placebo group**<br>**(N = 151)** | ***P* value** |
| --- | --- | --- | --- |
| Severe adverse events (at least one), n (%) | 9 (6.3) | 11 (7.3) | 0.819 |
| All-cause death, n (%) | 1 (0.7) | 3 (2.0) | 0.623 |
| Cardiovascular death, n (%) | 1 (0.7) | 2(1.3) | 1.000 |
| Non-cardiovascular death, n (%) | 0 (0.0) | 1(0.7) | 1.000 |
| Congestive heart failure, n (%) | 1 (0.7) | 4(2.6) | 0.371 |
| Cardiac shock, n (%) | 1 (0.7) | 1(0.7) | 1.000 |
| Cardiac rupture, n (%) | 0 (0.0) | 1(0.7) | 1.000 |
| Ventricular fibrillation, n (%) | 1 (0.7) | 2(1.3) | 1.000 |
| Stent thrombosis, n (%) | 0 (0.0) | 1 (0.7) | 1.000 |
| Angina, n (%) | 3 (2.1) | 0 (0.0) | 0.115 |
| Infectious pneumonia, n (%) | 0 (0.0) | 1 (0.7) | 1.000 |
| Stroke, n (%) | 1 (0.7) | 0 (0.0) | 0.488 |
| Respiratory failure, n (%) | 1 (0.7) | 0 (0.0) | 0.488 |
| Allergic shock, n (%) | 0 (0.0) | 1 (0.7) | 1.000 |
| Nephrotic syndrome, n (%) | 0 (0.0) | 1 (0.7) | 1.000 |
| Hypotension, n (%) | 0 (0.0) | 1 (0.7) | 1.000 |

## DISCUSSION

In this double-blind, placebo-controlled randomized clinical trial, intravenous Shenfu injection administered prior to reperfusion and followed by once a day until 5 days after primary PCI for STEMI did not reduce myocardial infarct size compared with placebo. There was also no effect on any of the secondary endpoints. The drug was safe and well tolerated.

Multiple mechanical and pharmacological strategies aimed at reducing infarct size after STEMI have failed^11, 22–27^. One major reason might be that key mechanisms involved in the pathophysiology of reperfusion injury are manifold and complex, all of which have been identified as therapeutic targets for inhibiting myocardial injury^23^. A single-target agent or intervention may be ineffective to cardioprotection against reperfusion injury. In this regard, emerging studies suggested that the adoption of multitargeted therapy directed to distinct targets may provide synergistic cardioprotective effects, and further achieve the optimal cardioprotection^28–31^.

As a Chinese herbal formula, Shenfu injection has shown satisfactory therapeutic efficacy in several other clinical setting including cardiogenic shock^32^, cardiac arrest^17^ and heart failure^33–35^, via the combination of multi-ingredients and multitargets. Extensive evidence from animal studies has indicated that Shenfu injection could effectively protect against sepsis-induced myocardial injury by inhibiting mitochondrial apoptosis^36^ and attenuating lipopolysaccharide-induced myocardial inflammation^37^, and it might regulate the expression of adenosine receptors to ameliorate the ischemia-reperfusion injury^38^. Moreover, our pilot study showed that the use of Shenfu in STEMI patients was safe and there was a trend towards reduction in CMR infarct size in the Shenfu injection group^20^. However, the RESTORE trial showed that there was no reduction of infarct size and other CMR parameters in patients receiving Shenfu injection. These findings further reconfirmed that reperfusion injury per se is a complex and multifactorial pathophysiological process, with multiple mechanisms.

Based on prior studies and position papers^2, 39, 40^, the RESTORE trial enrolled high-risk STEMI patients, presenting with proximal or mid LAD occlusion (and without prior myocardial infarction) prior to primary PCI (74.7% patients with pre-PCI TIMI flow 0 or 1 in the primary analysis population). The study population thus represents a highly selected cohort of patients with large infarcts, most likely to benefit from a novel cardioprotective therapy. Several possible factors may explain the lack of benefit of Shenfu injection in our study. First, one potential explanation may be an imbalance in age and pre-PCI TIMI flow at baseline between groups. Previous studies showed that age and pre-PCI TIMI flow grade 0 were the determinates of infarct size^41, 42^. Shenfu injection group had significantly higher rates of pre-PCI TIMI flow grade 0 in the primary analysis population (80.6% versus 69.1%; P = 0.036, Supplemental Table 3) compared with the placebo group. This discrepancy may contribute to the absence of the efficacy of Shenfu injection in reducing the infarct size. Second, potential benefit of Shenfu injection cannot be excluded in a certain subgroup of patients with severe hemodynamic changes (cardiogenic shock or those in Killip class III-IV), who have greater opportunity to obtain additional myocardial salvage, as well as those that are ineligible for CMR imaging. Third, no dose-response studies for assessment of the optimal dose and duration of Shenfu injection for myocardial infarction have been done, although the dosage and duration in this study were based on prior clinical data and were limited by the known side effects^20^. The optimal required concentration of Shenfu injection may be higher than were actually achieved in this study. And it is unclear whether or not the dose and concentration of Shenfu injection achieved is optimal at the time of reperfusion due to pharmacodynamic limitations. Future efforts should consider blood sample storage at different time points to provide deeper insights in optimal therapeutic concentrations. Finally, the failed translation from animal models to clinical efficacy in ameliorating myocardial injury might result from the absence of comorbidities and comedications in animal experiments.

Shenfu injection is a widely used drug with acceptable safety profiles. Metabolism and nutrition disorders (e.g. hypokalemia), hepatobiliary system disorders (e.g. liver injury) and gastrointestinal disorders (e.g. nausea) represent well-known, mostly mild side effects of Shenfu injection and were not more frequently reported in the Shenfu injection group.

### Limitations

This study has limitations. First, the present trial included anterior STEMI patients with proximal/mid LAD occlusion. Subjects with severe heart failure (Killip class III or above) were excluded. Thus, the study results might not be applicable to other sites of myocardial infarction or critically ill patients. Secondly, only a single dose and duration of Shenfu injection was evaluated in this trial. It is possible that the Shenfu dose regimen and duration were insufficient to achieve the optimal cardioprotective effect for STEMI patients. Third, the emergency of the national COVID-19 pandemic during the study period raised a big obstacle including pauses in enrolment due to lockdown, and limited completion of CMR imaging due to potential COVID-19 infection.

### Conclusions

Among patients with acute anterior STEMI presenting within 12 hours of symptoms, adjunctive Shenfu injection before reperfusion was safe but did not mitigate myocardial injury in the contemporary era.

## Data Availability

Data are not currently available. Will data be available: Yes Where: Electronic repository When will data availability begin: Date to be confirmed by the corresponding author.

## Acknowledgements

NA.

## Nonstandard Abbreviations and Acronyms

AE: adverse event
CMR: cardiac magnetic resonance
LAD: left anterior descending
LV: left ventricular
MACCE: major adverse cardiovascular and cerebrovascular event
PCI: percutaneous coronary intervention
SAE: serious adverse event
STEMI: ST-segment elevation myocardial infarction
TIMI: thrombolysis in myocardial infarction

